# Liver Biomarker Improves AHA/ACC 10-year ASCVD Risk Prediction in US and China Cohorts with ML

**DOI:** 10.64898/2026.04.22.26351466

**Authors:** Tongyu Peng, Chenglin Liu

## Abstract

**Introduction:** Accurate stratification of hard atherosclerotic cardiovascular disease (ASCVD) risk remains challenging despite advances in prevention. Liver function biomarkers (LFBs), particularly gamma-glutamyl transferase (GGT), have been linked to cardiovascular outcomes, yet their contribution to hard ASCVD risk prediction is not well defined.

**Methods:** This study analyzed data from the National Health and Nutrition Examination Survey (NHANES, 2005–2018) to assess cross-sectional associations between LFBs and 10-year hard ASCVD risk estimated by the ACC/AHA Pooled Cohort Equations. Multivariable regression, restricted cubic splines, and mediation analyses were applied to examine independent and dose–response relationships. External validation was performed in the China Health and Retirement Longitudinal Study (CHARLS) and NHANES using machine learning models (CoxBoost, Naive Bayes and Random Forest).

**Results:** Among 5,731 NHANES participants, GGT showed an independent linear association with hard ASCVD risk (P-trend = 0.003), partly mediated by systolic blood pressure (44.8%), HbA1c (19.0%), and high density lipoprotein cholesterol (13.4%). Machine learning (ML) models incorporating GGT, alkaline phosphatase (ALP), and globulin alongside traditional risk factors improved predictive accuracy, with Naive Bayes achieving an AUC of 0.751 in NHANES validation.

**Conclusions:** GGT is an independent and biologically plausible biomarker of hard ASCVD risk, acting through cardiometabolic pathways. Incorporating LFBs into risk prediction models, particularly with machine learning, enhances risk stratification and may facilitate early identification of high-risk individuals.

## 1. Introduction

Cardiovascular diseases (CVDs) persist as the foremost cause of global morbidity and mortality, posing a substantial public health challenge and socioeconomic burden[1,2]. Despite considerable progress in the prevention, diagnosis, and treatment of CVDs over recent decades, their overall burden remains substantial, and current risk stratification tools still exhibit limitations in predictive accuracy[3,4]. Therefore, identifying novel, readily accessible indicators with independent predictive value is essential for improving early CVD risk assessment, informing precision prevention strategies, and ultimately reducing the disease burden.

Accumulating evidence has established a strong link between CVD and abnormal metabolism, which can be quantified through serum biomarkers. Liver function is typically evaluated using a panel of routine biochemical markers, which has been shown to negatively affect both liver function and cardiovascular health[5–9]. Liver function biomarkers (LFBs) have emerged as potential predictors of cardiovascular risk. Previous studies have revealed complex associations between various liver function indicators and CVD risk[10,11]. For instance, γ-glutamyl transferase (GGT) has been identified as a risk factor for hard atherosclerotic cardiovascular disease (ASCVD)[12–14]. In contrast, the prognostic value of the aspartate aminotransferase to alanine aminotransferase (AST/ALT) ratio remains controversial. Labayen et al. reported that a lower AST/ALT ratio was associated with higher cardiometabolic risk factors[15], whereas research by Weng et al. suggested that an elevated AST/ALT ratio was significantly associated with an increased risk of CVD in men[16]. A multicenter retrospective study by Feng et al. also pointed out that peritoneal dialysis patients with higher AST/ALT ratios might face a significant risk of CVD mortality[17]. Although the precise impact of the AST/ALT ratio on CVD remains controversial, its abnormal variations are generally regarded as indicative signals of CVD risks. Additionally, hypoalbuminemia has been identified as an independent adverse prognostic indicator for CVD[18–20]. Collectively, these findings suggest that abnormalities in liver function indicators could serve as potential predictors of cardiovascular risk.

A substantial proportion of mortality in CVD is attributed to atherosclerosis. Hard ASCVD, defined as coronary death, nonfatal myocardial infarction, or fatal or nonfatal stroke, often manifests as sudden acute events, such as myocardial infarction, which are challenging to prevent. Traditional biomarkers, including creatine kinase-MM, creatine kinase-MB, and lactate dehydrogenase (LDH), have long been recognized as valuable diagnostic indicators for acute myocardial injury. However, these acute-phase biomarkers are not suitable for assessing the chronic progression or early detection of atherosclerosis. To facilitate prevention, well-established risk prediction models, such as the 10-year Hard ASCVD risk score developed by the American College of Cardiology/American Heart Association (ACC/AHA), have been widely applied[21]. Nevertheless, these conventional models primarily focus on lipid levels, glucose metabolism, and vascular stiffness, while largely neglecting the potential impact of metabolic abnormalities.

In this context, the present study aimed to refine ASCVD risk assessment by incorporating liver function enzymes, which may provide additional insights into the metabolic contributions to atherosclerosis. Leveraging data from the National Health and Nutrition Examination Survey (NHANES, 2005–2018), we evaluated the independent associations between a series of routinely measured liver function indicators and Hard ASCVD scores. Furthermore, the China Health and Retirement Longitudinal Study (CHARLS) was employed to validate the predictive performance of the 10-year Hard ASCVD risk estimator for clinical hard endpoints using machine learning approaches. The findings of this study are expected to provide novel epidemiological evidence on the role of liver-related biomarkers in cardiovascular health and contribute to optimizing risk assessment strategies for Hard ASCVD. This cross-sectional study has been reported in line with the STROCSS guidelines[22].

## 2. Materials and Methods

### 2.1 Study Design and Data Source

This cross-sectional study used data from seven consecutive NHANES cycles from 2005/2006 through 2017/2018. Detailed information on survey design, protocols, operational manuals, and publicly available datasets is available on the National Center for Health Statistics website (www.cdc.gov/nchs/nhanes/index.htm). The participant selection process is presented in the study flowchart (***Fig. 1A***). We included participants with available liver function biomarker data and an eligible Hard ASCVD risk score. We excluded participants who were pregnant or who had cardiovascular disease or cancer and then excluded participants with missing covariate data. Ultimately, 5,731 high quality participants aged 40 to 79 were retained. For machine learning study, CHARLS participants were extracted from waves 1 to 5 and had no cardiovascular disease at the 2011 baseline interview. Finally, 5,136 participants aged 45 to 79 with complete survival time and outcome data in the ten years follow-up were selected, as shown in ***Fig. 1B***.

**Figure 1.**
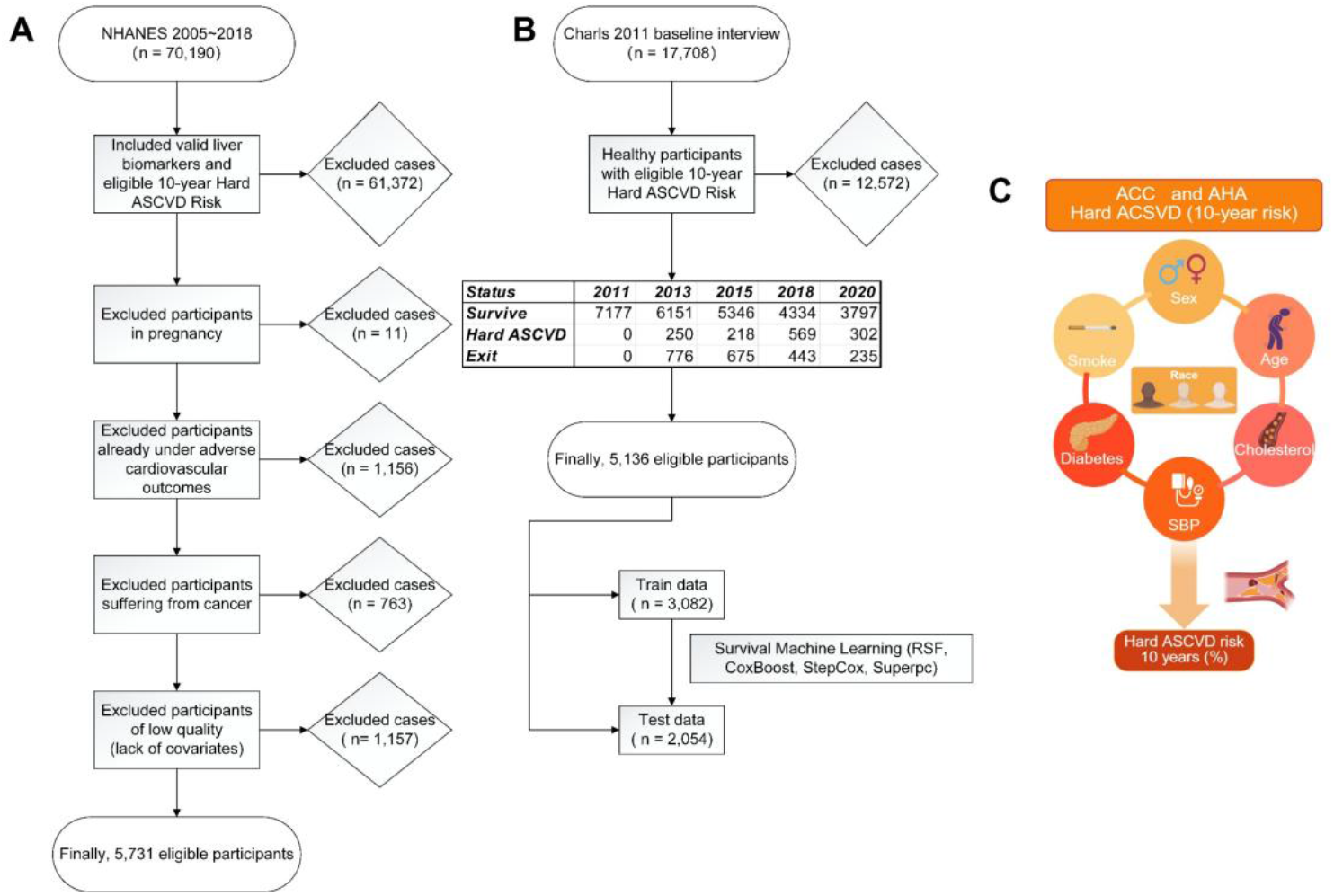
Flow chart of data processing and study design. (A) NHANES data processing details. (B) Charls data processing details. (C) Components of the ACC/AHA Hard ASCVD ten-year risk score.

### 2.2 Exposure variables: LFBs

The LFBs included GGT (IU/L), alkaline phosphatase (ALP, IU/L), aspartate aminotransferase (AST, IU/L), alanine aminotransferase (ALT, IU/L), LDH (IU/L), albumin (g/dL), and globulin (g/dL). The AST to ALT ratio and the albumin to globulin ratio (A/G ratio) were calculated.

### 2.3 Outcome Variable: Hard ASCVD risk calculation

This study used the Pooled Cohort Equations recommended in the 2013 American College of Cardiology/American Heart Association (ACC/AHA) guidelines to estimate the ten year risk of a first hard ASCVD event[21]. The variables required to estimate ASCVD risk were age, sex, race, total cholesterol, high density lipoprotein cholesterol (HDL-C), systolic blood pressure, use of antihypertensive medication, diabetes status, and smoking status (***Fig*.*1C***). Consistent with clinical guidelines, a risk scores greater than 7.5% was classified as high risk. The Hard ASCVD risk was calculated using four equations (1-1), (1-2), (1-3), and (1-4). Detailed variables and coefficients are provided in ***Supplementary Table 1***.

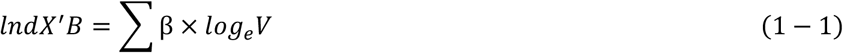

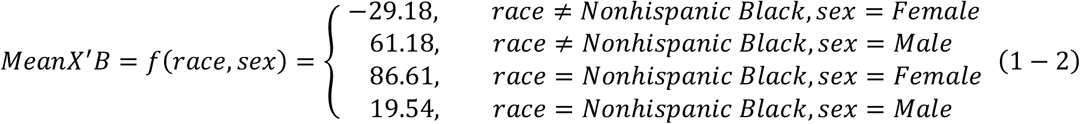

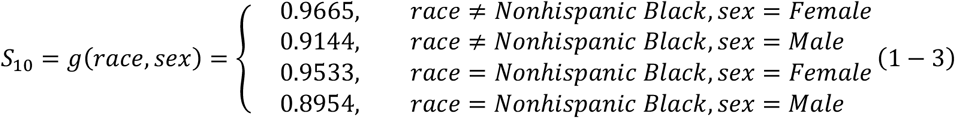

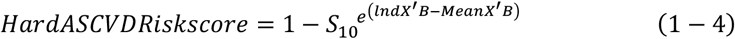

### 2.4 Covariates and definitions

Sociodemographic variables included age, sex, race, poverty to income ratio (PIR), and educational attainment. Lifestyle factors comprised smoking status and alcohol consumption. Health-related variables included diabetes (defined as glycosylated hemoglobin A1c [HbA1c] ≥ 6.5%, use of glucose lowering medication, or a prior physician diagnosis) and hypertension (defined as systolic blood pressure [SBP] ≥ 140 mmHg, diastolic blood pressure [DBP] ≥ 90 mmHg, or use of antihypertensive medication). Serum cholesterol (total cholesterol, HDL-C and low density lipoprotein cholesterol [LDL-C]) was also recorded.

### 2.5 Statistical analysis for cross-sectional associations

LFBs were categorized into tertiles, and indicator variables for tertile membership were created. Trend analyses across tertiles were performed using three models, and P for trend was calculated. The models were specified as follows: Crude model (no covariate adjustment); Multivariable model 1 (adjusted for sociodemographic variables: sex, age, race/ethnicity, marital status, poverty to income ratio, and educational attainment); and Multivariable model 2 (additionally adjusted for smoking status, hypertension status, diabetes status, and HDL-C). To investigate dose response relations, restricted cubic splines (RCS) were fitted. Mediation analyses were performed by mediation R package to examine potential mediating effects between LFBs and traditional risk factors (lipids, glucose, and blood pressure)[23].

### 2.6 Validation in cohort and machine learning approaches

To evaluate model performance against clinically ascertained hard ASCVD outcomes (coronary death, nonfatal myocardial infarction, or fatal or nonfatal stroke), CHARLS Hard ASCVD risk scores were calculated using the same equations applied to NHANES. Survival machine learning models were constructed using the Mime1 R package, which integrates multiple algorithms for prognostic modeling[24]. These models used the risk calculator features to predict observed clinical outcomes; Kaplan Meier curves were plotted to compare survival time and event status between high risk and low risk groups. Next, robust LFBs selected by LASSO and logistic regression were used to build binary classification models[25]. For both types of machine learning, CHARLS was split with 60% used for training and 40% used for testing, and the binary classification models were externally validated using NHANES. Because CHARLS did not include measured LFBs at the visit, missing LFB values were imputed with the k-Nearest Neighbor (kNN) function in the VIM R package; sample similarity was computed using a Gower-type distance based on shared variables (glycaemia, blood pressure, lipids, and demographic variables), and the two nearest donor samples were used for imputation[26,27]. This procedure allowed us to approximate the most likely liver function biomarker values for CHARLS participants, conditional on the covariate structure observed in NHANES.

All analyses were conducted using R software (version 4.3.3). The NHANES complex sampling design was consistently incorporated across all analyses. All statistical tests were two sided, with a significance threshold of P < 0.05.

## 3. Results

### 3.1 Cross-sectional Analysis

#### 3.1.1 Baseline Characteristics of Study Participants

A total of 5,731 eligible participants were included in the analysis. Participants were stratified according to the Hard ASCVD risk score. Significant differences were observed among groups with respect to sociodemographic characteristics, lifestyle factors, and clinical indicators. Overall, participants with higher Hard ASCVD risk scores tended to be older, were more likely to be male, and had higher prevalence rates of diabetes and hypertension. For LFBs, baseline descriptions indicated significant group differences in multiple serum enzymes and proteins, including GGT, AST/ALT ratio, ALP, LDH, A/G ratio, and globulin. Most of these differences were statistically significant (P < 0.001). Detailed means and P values are presented in ***Supplementary Table 2***.

#### 3.1.2 Multivariable Logistic Regression and Trend Analysis

To assess the independent associations between various liver function indicators and hard ASCVD risk, multivariable logistic regression analysis was employed, with the results presented in **Table 1**. Multivariate model 2, elevated serum GGT levels, maintained a significant independent positive association with hard ASCVD risk. T_2_ (OR=1.50, 95%CI=1.03 to 2.19) and T_3_ (OR=1.81, 95%CI=1.28 to 2.25) of GGT showed significant positive relationship of hard ASCVD risk (p for trend=0.003^**^). Although most LFBs exhibited significant relationship in hard ASCVD risk, they couldn’t maintain their performance when more variables were adjusted in model 1 and 2. In summary, the multivariable-adjusted analyses highlight the potential importance of GGT, as independent factors associated with hard ASCVD risk, and suggest that GGT may exhibit dose-response characteristics.

**Table 1.** Tendency analysis of liver function biomarkers in 3 models.

| **Biomarkers** | **Low**<br>**Risk** | **High**<br>**Risk** | **Crude OR**<br>**(95% CI)** | **Multivariate**<br>**model 1**<br>**OR (95% CI)** | **Multivariate**<br>**model 2**<br>**OR (95% CI)** |
| --- | --- | --- | --- | --- | --- |
| **ALP, IU/L** |  |  |  |  |  |
| T<sub>3</sub>>77 | 1023 | 845 | 1.43(1.21,1.69) | 1.47(1.13,1.91) | 0.76(0.53,1.07) |
| T<sub>2</sub> 60-77 | 1080 | 801 | 1.16(0.96,1.41) | 1.27(0.98,1.65) | 1.06(0.73,1.55) |
| T<sub>1</sub><60 | 1230 | 752 | Ref | Ref | Ref |
| *P for trend* |  |  | <0.001*** | 0.005** | 0.110 |
| **ALT, IU/L** |  |  |  |  |  |
| T<sub>3</sub>>26 | 1036 | 699 | 1.07(0.87,1.32) | 1.25(0.91,1.71) | 1.19(0.78,1.83) |
| T<sub>2</sub> 18-26 | 1117 | 868 | 1.16(0.99,1.36) | 0.98(0.77,1.24) | 1.20(0.86,1.67) |
| T<sub>1</sub><18 | 1180 | 831 | Ref | Ref | Ref |
| *P for trend* |  |  | 0.639 | 0.115 | 0.488 |
| **LDH, IU/L** |  |  |  |  |  |
| T<sub>3</sub>>142 | 963 | 912 | 1.63(1.30,2.03) | 0.74(0.55,1.00) | 0.85(0.58,1.25) |
| T<sub>2</sub> 121-142 | 1114 | 733 | 1.09(0.90,1.31) | 0.70(0.53,0.94) | 0.88(0.60,1.27) |
| T<sub>1</sub><121 | 1256 | 753 | Ref | Ref | Ref |
| *P for trend* |  |  | <0.001*** | 0.077 | 0.437 |
| **GGT, IU/L** |  |  |  |  |  |
| T<sub>3</sub>>27 | 978 | 882 | 1.95(1.63,2.33) | 2.57(1.91,3.46) | 1.81(1.28,2.55) |
| T<sub>2</sub> 17-27 | 1020 | 834 | 1.56(1.29,1.89) | 1.38(1.06,1.80) | 1.50(1.03,2.19) |
| T<sub>1</sub><17 | 1335 | 682 | Ref | Ref | Ref |
| *P for trend* |  |  | <0.001*** | <0.001*** | 0.003** |
| **AST, IU/L** |  |  |  |  |  |
| T<sub>3</sub>>26 | 982 | 711 | 1.22(0.99,1.51) | 0.84(0.61,1.15) | 0.74(0.49,1.10) |
| T<sub>2</sub> 21-26 | 972 | 776 | 1.26(1.04,1.52) | 0.91(0.68,1.22) | 1.20(0.80,1.80) |
| T<sub>1</sub><21 | 1379 | 911 | Ref | Ref | Ref |
| *P for trend* |  |  | 0.061 | 0.300 | 0.106 |
| **Albumin, g/dL** |  |  |  |  |  |
| T<sub>3</sub>>4.3 | 1050 | 684 | 0.81(0.66,0.98) | 0.84(0.66,1.07) | 1.06(0.73,1.53) |
| T<sub>2</sub> 4.1-4.3 | 873 | 641 | 0.99(0.81,1.22) | 1.00(0.74,1.35) | 1.30(0.94,1.80) |
| T<sub>1</sub><4.1 | 1410 | 1073 | Ref | Ref | Ref |
| *P for trend* |  |  | 0.034* | 0.156 | 0.849 |
| **Globulin, g/dL** |  |  |  |  |  |
| T<sub>3</sub>>3.1 | 905 | 796 | 1.56(1.30,1.86) | 1.82(1.34,2.47) | 1.28(0.88,1.87) |
| T<sub>2</sub> 2.7-3.1 | 1186 | 884 | 1.28(1.10,1.50) | 1.31(1.02,1.69) | 1.31(0.92,1.88) |
| T<sub>1</sub><2.7 | 1242 | 718 | Ref | Ref | Ref |
| *P for trend* |  |  | <0.001*** | <0.001*** | 0.122 |
| **AST/ALT** |  |  |  |  |  |
| T<sub>3</sub>>1.19 | 1055 | 855 | 1.16(0.96,1.41) | 0.53(0.39,0.72) | 0.74(0.48,1.12) |
| T<sub>2</sub> 0.95-1.19 | 1114 | 794 | 1.04(0.86,1.26) | 0.57(0.44,0.73) | 0.75(0.52,1.09) |
| T<sub>1</sub><0.95 | 1164 | 749 | Ref | Ref | Ref |
| *P for trend* |  |  | 0.111 | <0.001*** | 0.165 |

| A/G |  |  |  |  |  |
| --- | --- | --- | --- | --- | --- |
| T<sub>3</sub>>1.56 | 1163 | 697 | 0.71(0.59,0.86) | 0.68(0.51,0.92) | 1.13(0.75,1.70) |
| T<sub>2</sub> 1.33-1.56 | 1096 | 797 | 0.86(0.71,1.05) | 0.89(0.66,1.20) | 1.37(0.93,2.02) |
| T<sub>1</sub><1.33 | 1074 | 904 | Ref | Ref | Ref |
| *P for trend* |  |  | <0.001*** | 0.010* | 0.744 |

Crude model (no covariate adjustment); Multivariable model 1 (adjusted for sociodemographic variables: sex, age, race/ethnicity, marital status, poverty to income ratio, and educational attainment); and Multivariable model 2 (additionally adjusted for smoking status, hypertension status, diabetes status, and HDL-C). ALP, alkaline phosphatase; ALT, alanine aminotransferase; LDH, lactate dehydrogenase; GGT, -glutamyl transpeptidase; AST, aspartate transaminase; A/G, albumin / globulin. T<sub>1</sub>, the first tertile; T<sub>2</sub>, the second tertile; T<sub>3</sub>, the third tertile. \*, P< 0.05; \*\*, P < 0.01; \*\*\*, P < 0.001.

The RCS regression models were constructed after adjusting for the same covariates as in Multivariate model 2. The RCS analysis revealed a significant positive association between GGT and Hard ASCVD risk (P-overall < 0.001^***^). The non-linearity test was not significant (P = 0.3244), indicating that the relationship was effectively linear, as shown in ***Fig. 2***. The significant linearity of GGT was paralleled along with the results of multivariate models. Higher concentration of GGT in serum indicated higher risk of hard ASCVD.

**Figure 2.**
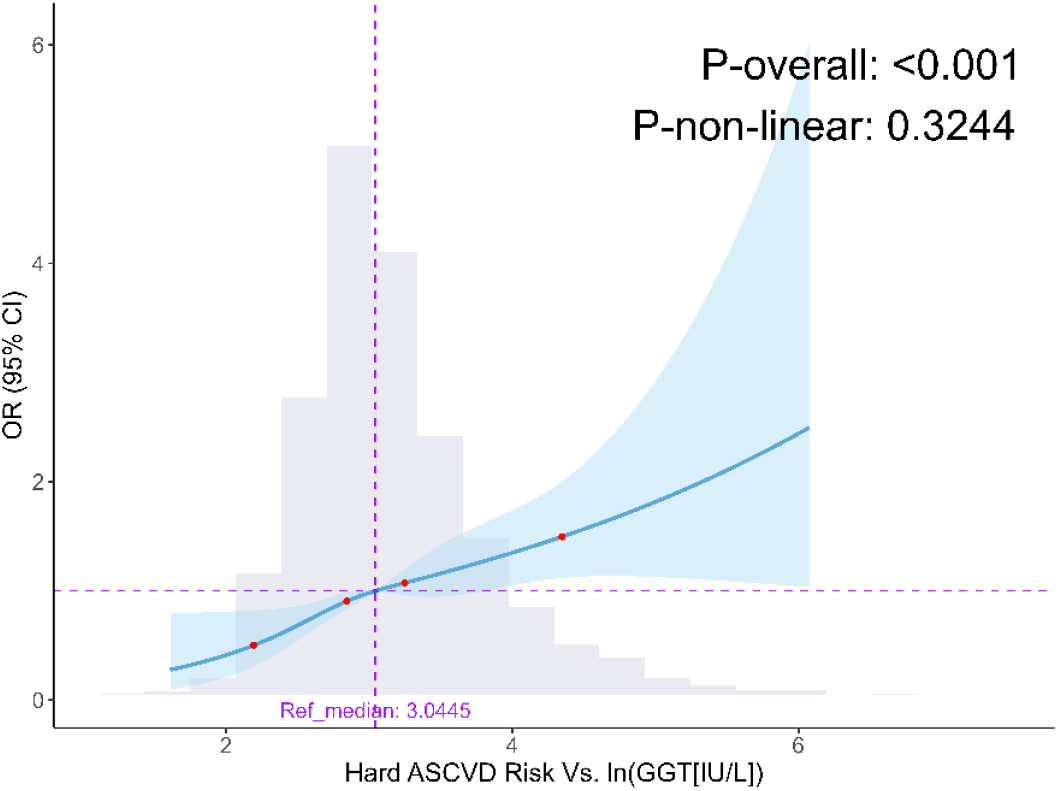
Restricted cubic spline (RCS) plots for the association between GGT and Hard ASCVD risk. The median value of GGT was used as the reference point. The vertical pink dashed line indicates the median, and the gray histogram shows the distribution of GGT. The blue curve depicts the odds ratio (OR) with changes in GGT, while the shaded area represents the 95% confidence interval (CI). Covariates were adjusted according to multivariate model 2.

#### 3.1.3 Mediation Effect of GGT

We examined whether HDL, HbA1c, and systolic blood pressure (SBP) mediated the association between serum GGT level and the Hard ASCVD risk score, as shown in ***Fig. 3***. All examined path coefficients were highly significant (p < 0.001). Specifically, higher GGT was associated with lower HDL (β = −1.2732, p < 0.001), and HDL was negatively associated with the Hard ASCVD risk score (β = −0.0477, p < 0.001). In the HDL-mediated model the indirect effect (IE) was 0.0038 (95% CI: 0.0010 to 0.0071, p < 0.001) and the direct effect (DE) was 0.0250 (95% CI: 0.0132 to 0.0348, p < 0.001), with the mediation proportion ≈ **13.4%**, indicating a partial mediation by HDL.

**Figure 3.**
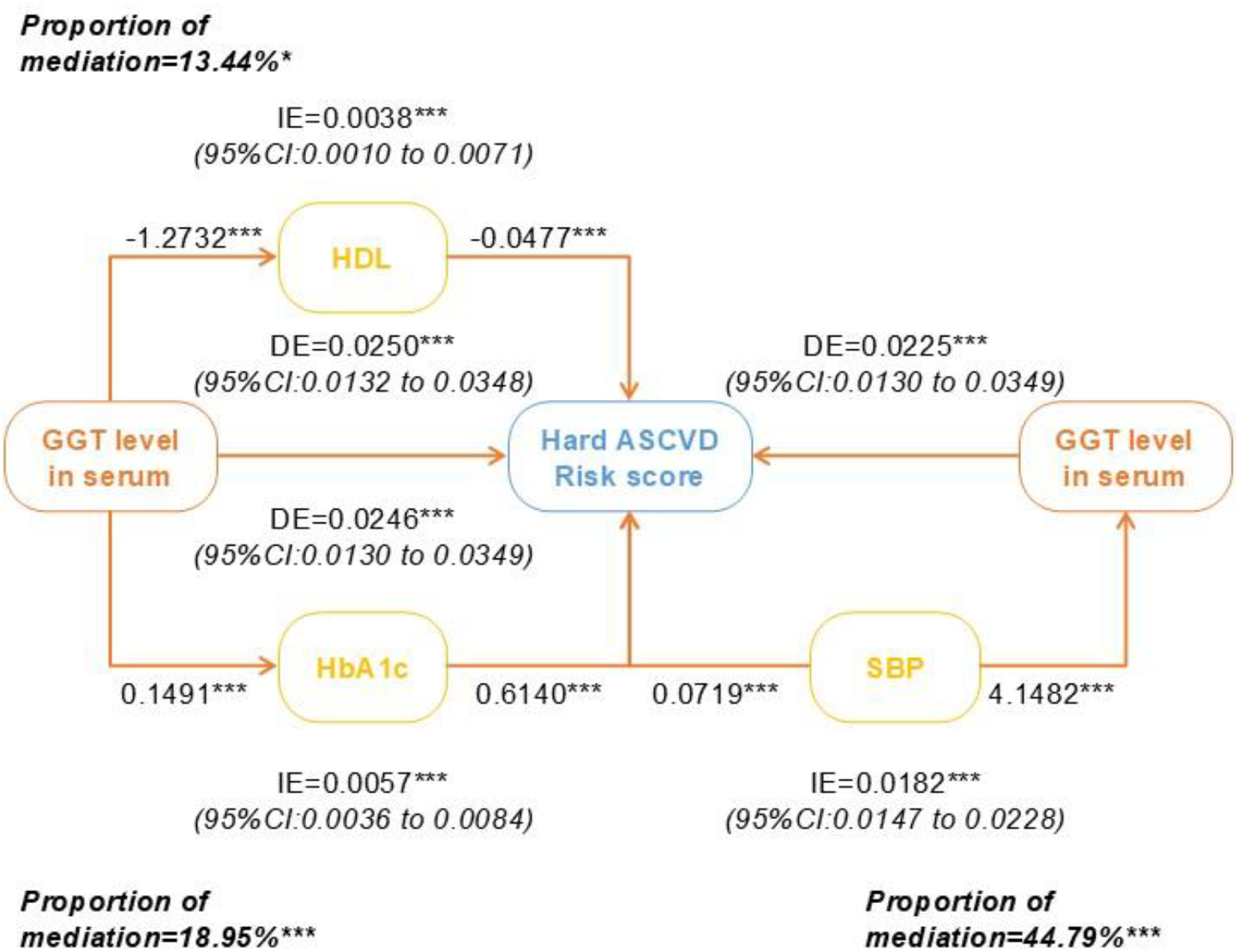
Mediation effect of GGT. IE, indirect effect; DE, direct effect. ^*^ p<0.05, ^**^p<0.01, ^***^p<0.001

In the HbA1c-mediated model, GGT was positively associated with HbA1c (β = 0.1491, p < 0.001), and HbA1c was positively associated with the risk score (β = 0.6140, p < 0.001). The indirect effect was 0.0057 (95% CI: 0.0036 to 0.0084, p < 0.001) and the direct effect was 0.0246 (95% CI: 0.0130 to 0.0349, p < 0.001); the mediation proportion was approximately **19.0%**, indicating that glucose metabolism (HbA1c) accounted for a meaningful portion of the association.

In the SBP-mediated model, GGT was strongly positively associated with SBP (β = 4.1482, p < 0.001), and SBP was positively associated with the risk score (β = 0.0719, p < 0.001). The indirect effect via SBP was 0.0182 (95% CI: 0.0147 to 0.0228, p < 0.001) while the direct effect was 0.0225 (95% CI: 0.0130 to 0.0349, p < 0.001). SBP accounted for the largest mediation proportion (∼**44.8%**), indicating that blood pressure is a major pathway linking GGT to Hard ASCVD risk.

Collectively, these results indicate that serum GGT relates to Hard ASCVD risk both directly and indirectly through HDL, glycemic control (HbA1c), and especially SBP. All models were adjusted for the covariates described in the Methods.

### 3.2 Machine learning

#### 3.2.1 Calculator validation and Survival machine learning

To validate the clinical utility of the traditional 10-year Hard ASCVD risk calculator, 5,136 CHARLS participants with clearly defined outcomes during the 10 years follow up were included. The baseline year was 2011 (CHARLS wave 1), and risk scores were calculated using baseline data. Scores were log-transformed to approximate a normal distribution, and survival models were fitted with log transformed risk scores and survival time.

As shown in ***Fig. 4A***, restricted cubic spline analysis demonstrated a positive linear association between predicted risk scores and observed hard ASCVD outcomes. Across both low risk and high risk strata, higher risk scores were consistently associated with increased hazard ratios (HR). Participants classified as high risk (risk score > 7.5%) exhibited significantly higher HR compared with those in the low risk group.

**Figure 4.**
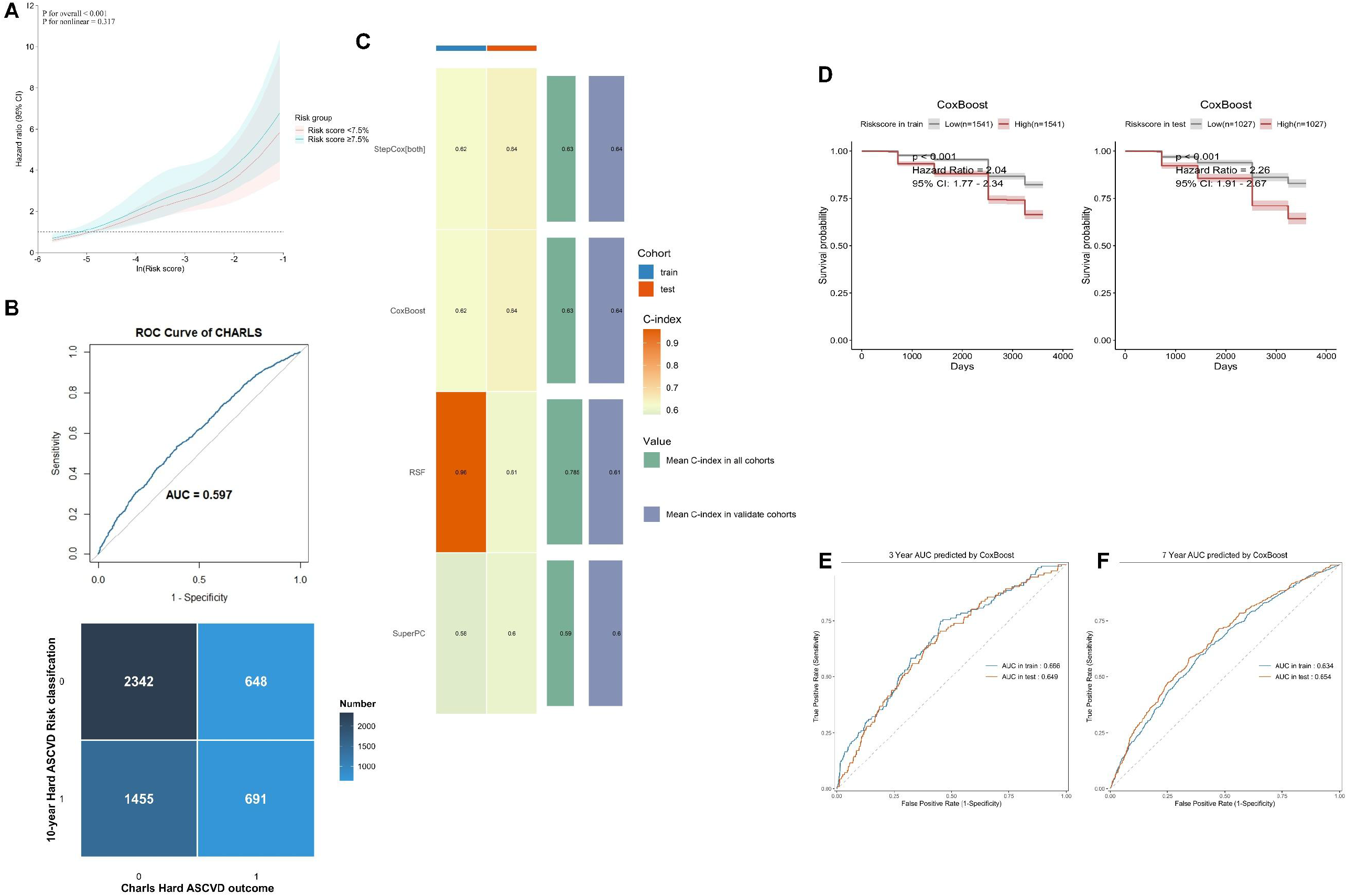
Clinical validations of the 10-year Hard ASCVD risk calculator. (A) Restricted cubic spline (RCS) plot of log transformed risk score and CHARLS outcomes. (B) ROC curve of risk score in CHARLS dataset; confusion matrix comparing predicted Hard ASCVD risk categories with observed outcomes in CHARLS. (C) Performance of survival machine learning models using traditional risk calculator features. (D) Kaplan–Meier survival curves of training and test datasets in the CoxBoost algorithm. (E, F) Receiver operating characteristic (ROC) curves of CoxBoost at 3 years (E) and 7 years (F).

Nevertheless, the predictive accuracy of the traditional calculator was modest. As shown in ***Fig. 4B***, the ROC curve showed that the risk score achieved an AUC of 0.597 in the CHARLS dataset judging the hard ASCVD condition. And the confusion matrix indicated an overall accuracy of 59.0% for discriminating against clinical outcomes.

To further investigate predictive performance, survival machine learning approaches were applied. The features were the same as the features of AHA/ACC hard ASCVD calculator. And the outcome variable was clinical hard ASCVD first discovery. As presented in ***Fig. 4C***, CoxBoost and StepCox achieved superior discrimination compared with RSF and SuperPC, with mean C indices of 0.64 in validation cohorts. Survival curves (***Fig. 4D***) further confirmed that participants in the high risk group had significantly shorter survival times than those in the low risk group, with HRs exceeding 2.0 in both training and testing sets (both P < 0.001). And the C index was higher than 59% compared to the AHA/ACC risk calculator.

Time dependent ROC analyses (***Fig. 4E–F***) showed that CoxBoost maintained good discrimination over time, with AUCs of 0.656 and 0.649 at 3 years, and 0.634 and 0.654 at 7 years, in training and testing cohorts, respectively. These findings indicate that while the traditional calculator provides only moderate accuracy, machine learning models such as CoxBoost offer improved risk stratification and survival prediction.

#### 3.2.2 Machine learning including LFBs

As shown in Fig. 5A, among 5,136 CHARLS participants, 50 cases without body measure index (BMI) data were excluded, leaving 5,086 samples for analysis. This exclusion was necessary because BMI was one of the predictors identified by Least Absolute Shrinkage and Selection Operator (LASSO) algorithm. Feature selection using the LASSO algorithm yielded a parsimonious set of variables, including age, sex, HDL cholesterol, mean systolic blood pressure, ALP, GGT, smoking status, non-HDL cholesterol, antihypertensive treatment, HbA1c percentage, BMI, triglycerides, globulin, and alcohol consumption. The optimal penalty parameter was determined at the lambda 1 standard error threshold (***Fig. 5B***).

**Figure 5.**
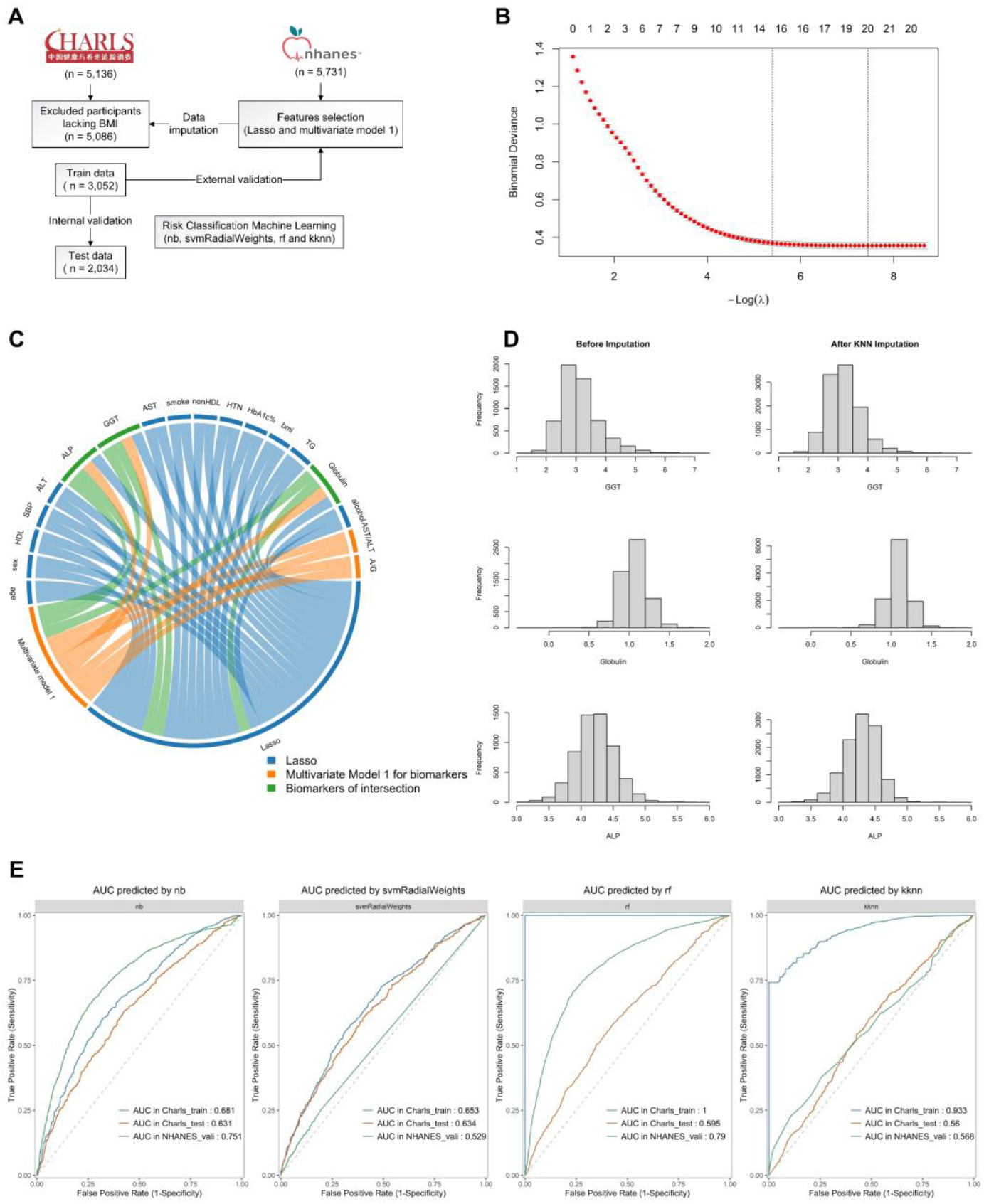
Risk classification machine learning. (A) A flowchart of machine learning; (B) feature selected by Lasso algorithm, the first dashed line representing lambda 1 SE; (C) Chord diagram showing the common biomarkers in both Lasso and Multivariate model 1; (D) Distribution of GGT, ALP and Globulin in Charls and NHANES; (E) ROC curves of machine learning in Naive Bayes (nb), Support Vector Machine (svmRadialWeights), random forest (rf) and K-nearest Neighbors(kknn).

Regarding LFBs, the chord diagram (***Fig. 5C***) highlights overlapping predictors identified by both LASSO and Multivariate Model 1, emphasizing GGT, ALP, and globulin as robust biomarkers. Following kNN imputation, the distributions of these biomarkers in CHARLS were comparable with those in NHANES, supporting the validity of the imputation strategy (***Fig. 5D***).

To evaluate predictive performance, multiple machine learning algorithms were applied. In CHARLS, the outcome was defined as the first occurrence of hard ASCVD or fatal hard ASCVD during 10 years of follow up. In NHANES, outcomes were defined according to high versus low risk classification based on the 7.5% threshold from the original risk calculator. This reverse validation using clinical outcomes provides a more rigorous evaluation of the incremental value of incorporating LFBs into prediction models.

As shown in ***Fig. 5E***, Naive Bayes, Support Vector Machine, Random Forest, and kNN models all demonstrated reasonable discrimination, with area under the curve (AUC) values consistently exceeding 0.6. Among them, Naive Bayes achieved the best overall performance, with AUCs of 0.681 in CHARLS training data, 0.631 in CHARLS test data, and 0.751 in NHANES external validation, indicating stable though moderate predictive accuracy across cohorts.

## 4. Discussion

In this large cross-sectional study using nationally representative NHANES data, we found that serum GGT levels were independently, positively, and linearly associated with the 10-year risk of hard ASCVD, as estimated by the ACC/AHA Pooled Cohort Equations. This relationship showed a clear dose-response pattern and remained robust after adjusting for extensive demographic, lifestyle, and clinical confounders. Mediation analysis further indicated that systolic blood pressure, HbA1c, and HDL-C accounted for substantial proportions of this association. Importantly, external validation with NHANES demonstrated that incorporating selected LFBs (GGT, ALP, and globulin) alongside traditional risk factors significantly improved the predictive performance of machine learning models for incident ASCVD events.

These results corroborate prior epidemiological studies identifying GGT as a robust independent predictor of kinds of CVDs and all-cause as well as cardiovascular mortality[13,28–32]. The novel contribution of this study is the demonstration of GGT’s independent association with the comprehensive hard ASCVD risk score recommended by clinical guidelines. The restricted cubic spline analysis confirmed a linear dose-response, indicating that even modest increases in GGT may translate into graded cardiovascular risk without a threshold effect. This supports the role of GGT as a continuous risk marker.

Mediation analysis highlighted systolic blood pressure as the dominant pathway, accounting for ∼45% of the GGT–ASCVD association. This aligns with the role of GGT in glutathione metabolism and oxidative stress, which impair nitric oxide bioavailability and promote vascular dysfunction[33,34]. Cheraghi M, et al, directly validated positive assortation between GGT and oxidative stress levels in coronary heart disease cohorts[35]. HbA1c (19%) and HDL-C (13%) were also significant mediators, suggesting links to insulin resistance, dyslipidemia, and hepatic steatosis[36–38]. GGT was confirmed to be the independent risk factor of insulin resistance in steatotic patients[39]. Collectively, these results imply that GGT may participate in central pathophysiological processes (hyperglycosemia, dyslipidemia and hypertension) underlying ASCVD, beyond being a bystander biomarker.

A major strength of this study lies in the validation using the CHARLS cohort. While the ACC/AHA risk calculator showed modest discrimination (AUC of 0.597), survival machine learning models achieved better performance (C-index ∼0.64). Further integration of liver biomarkers, consistently identified by both LASSO and traditional models, enhanced predictive accuracy, with the Naive Bayes classifier yielding an AUC of 0.751 in the NHANES validation set. These findings underscore the incremental predictive value of liver biomarkers and support their generalizability across diverse populations.

Several limitations should be acknowledged. First, the cross-sectional design of NHANES precludes causal inference. Second, single measurements of liver enzymes may not capture long-term variability. Third, residual confounding by factors such as inflammation, genetics, or diet cannot be ruled out. Fourth, imputation of missing liver biomarkers in CHARLS, although performed with a robust kNN approach, introduces uncertainty. Lastly, while CHARLS provided clinical endpoints, NHANES analyses relied on calculated risk scores rather than observed events.

This study provides strong evidence that GGT is an independent and biologically plausible biomarker of hard ASCVD risk, acting through blood pressure, glycemic, and lipid pathways. Incorporating LFBs, particularly GGT, into existing risk assessment frameworks—augmented by machine learning—could improve early identification of high-risk individuals. As liver function tests are routine, inexpensive, and widely available, their inclusion in cardiovascular prevention strategies represents a pragmatic step toward more precise and personalized risk prediction. Future prospective studies are needed to confirm these findings and refine updated risk equations.

## 5. Conclusions

In this study, using cross-sectional data from NHANES, we demonstrated that GGT remains an independent and significant risk factor for hard ASCVD, even after adjustment for multiple covariates, exhibiting a robust linear association. Mediation analysis further revealed that the impact of GGT on hard ASCVD risk is significantly mediated through elevations in HbA1c% and SBP, as well as reductions in HDL-C. Subsequent survival machine learning models constructed from five waves of CHARLS observational data showed superior predictive accuracy compared with traditional AHA/ACC risk equations. Incorporating LFBs into binary prediction models further enhanced performance, achieving strong predictive capability in both CHARLS and external NHANES validation cohorts. Elevated GGT may serve as an enhancer of cardiovascular risk, and future multicenter studies developing risk equations should consider including robust LFBs such as GGT. Ultimately, liver function markers may provide valuable insights for early clinical prevention and screening of hard ASCVD.

## Data Availability

NHANES datasets are available on the National Center for Health Statistics website (www.cdc.gov/nchs/nhanes/index.htm). CHARLS datasets can be found at https://charls.pku.edu.cn/. Coding manuscripts and filtered datasets are available under request. And detailed coding manuscripts could be found at GitHub: CHENGlinLiuleen/ASCVDvalidation.

https://github.com/CHENGlinLiuleen/ASCVDvlidation

https://www.cdc.gov/nchs/nhanes/index.htm

https://charls.pku.edu.cn/

## Acknowledgements

We thank all the members of this research as well as all participants of CHARLS and NHANES for their contribution.

