## Supplement Table 1 for "Liver Biomarker Improves AHA/ACC 10-year ASCVD Risk Prediction in US and China Cohorts with ML"

Table S1. Detailed variables of hard ASCVD calculator

| Variables | White and other races | Non-Hispanic black |
| --- | --- | --- |
|  | Coefficient | Coefficient |
| Female | | |
| Ln Age | -29,799 | 17.114 |
| (Ln Age)^2^ | 4.884 | NA |
| Ln Total Cholesterol (mg/dL) | 13.540 | 0.94 |
| Ln Age x Ln Total cholesterol | -3.114 | NA |
| Ln HDL-C (mg/dL) | -13.578 | -18.920 |
| Ln Age x Ln HDL-C | 3.149 | 4.475 |
| Ln Treated SBP (mmHg) | 2.019 | 29.291 |
| Ln Age x Ln Treated SBP | NA | -6.432 |
| Ln Untreated SBP (mmHg) | 1.957 | 27.820 |
| Ln Age x Ln untreated SBP | NA | -6.807 |
| Ln Current Smoker (Yes=e, No=1) | 7.574 | 0.691 |
| Ln Age x Ln current smoker | -1.665 | NA |
| Ln diabetes (Yes=e, No=1) | 0.661 | 0.874 |
| Male | | |
| Ln Age | 12.344 | 2.469 |
| Ln Total Cholesterol (mg/dL) | 11.853 | 0.302 |
| Ln Age x Ln Total cholesterol | -2.664 | NA |
| Ln HDL-C (mg/dL) | -7.990 | -0.307 |
| Ln Age x Ln HDL-C | 1.769 | NA |
| Ln Treated SBP (mmHg) | 1.797 | 1.916 |
| Ln Untreated SBP (mmHg) | 1.764 | 1.809 |
| Ln Current Smoker (Yes=e, No=1) | 7.837 | 0.549 |
| Ln Age x Ln current smoker | -1.795 | NA |
| Ln diabetes (Yes=e, No=1) | 0.658 | 0.645 |

HDL-C, high density lipoprotein cholesterol; DBP, diastolic blood pressure; SBP, systolic blood pressure. Samples participated in this formula are whose low density lipoprotein cholesterol is <190 mg/dL.
