## Supplement Table 2 for "Liver Biomarker Improves AHA/ACC 10-year ASCVD Risk Prediction in US and China Cohorts with ML"

Table S2. Baseline Characteristics of NHANES

| Variables | level | Low Risk | High Risk | p |
| --- | --- | --- | --- | --- |
| n |  | 3333 | 2398 |  |
| Age (year) |  | 50.14 (6.89) | 63.18 (8.73) | <0.001^***^ |
| Sex | Female | 2013 (57.6%) | 906 (36.6%) | <0.001^***^ |
|  | Male | 1320(42.4%) | 1492 (63.4%) |  |
| Race | Mexican American | 580 (7.3%) | 358 (5.6%) | <0.001^***^ |
|  | Non-Hispanic Asian | 255 (2.6%) | 146 (2.8%) |  |
|  | Non-Hispanic Black | 519 (7.2%) | 644(13.3%) |  |
|  | Non-Hispanic White | 1503(74.2%) | 925 (70.2%) |  |
|  | Other Hispanic | 340 (4.5%) | 259 (4.9%) |  |
|  | Other Race - Including Multi-Racial | 136 (4.2%) | 66 (3.1%) |  |
| Marital | Married | 2232 (70.4%) | 1486(68.1%) | 0.223 |
|  | Unmarried | 1101 (29.6%) | 912 (31.9%) |  |
| Education Level | 9-11th grade | 408(8.8%) | 391 (11.7%) | <0.001^***^ |
|  | College graduate or above | 987 (35.9%) | 447 (23.4%) |  |
|  | High school graduate/GED or equivalent | 668 (20.1%) | 617(28.9%) |  |
|  | Less than 9th grade | 290 (4.2%) | 335 (7.4%) |  |
|  | Some college or AA degree | 980 (31.0%) | 608 (28.6%) |  |
| Poverty-to-income Ratio |  | 3.44 (1.58) | 3.02 (1.60) | <0.001^***^ |
| Body Measure Index (kg/m²) |  | 29.15 (6.64) | 30.04 (6.61) | 0.001^**^ |
| Drinking History | NO | 1046 (25.0) | 795 (27.6) | 0.159 |
|  | YES | 2287 (75.0) | 1603 (72.4) |  |
| Smoking Status | Current smoke | 496 (14.1%) | 620 (27.2) | <0.001^***^ |
|  | Never smoke | 2009 (58.5%) | 1029 (41.7%) |  |
|  | Quit <1y | 63 (2.1%) | 33(1.4%) |  |
|  | Quit >5y | 661 (22.0%) | 655(27.0%) |  |
|  | Quit 1~5y | 104 (3.2%) | 61 (2.7%) |  |
| Diabetes | NO | 3083 (94.1%) | 1623 (72.0%) | <0.001^***^ |
|  | YES | 250 (5.9%) | 775 (28.0%) |  |
| DBP (mmHg) |  | 72.13 (10.05) | 71.40 (13.43) | 0.102 |
| SBP (mmHg) |  | 118.81 (13.30) | 133.44 (17.18) | <0.001^***^ |
| Hypertension | NO | 2425 (72.4%) | 748 (33.8%) | <0.001^***^ |
|  | YES | 908 (27.6%) | 1650 (66.2%) |  |
| Total Cholesterol (mg/dL) |  | 199.57 (32.74) | 198.47 (35.61) | 0.443 |
| Non-HDL-C (mg/dL) |  | 143.07 (32.34) | 145.95 (36.12) | 0.063 |
| HDL-C (mg/dL) |  | 56.50 (14.86) | 52.52 (14.63) | <0.001^***^ |
| ALP (IU/L) |  | 67.55 (21.27) | 71.80 (24.19) | <0.001^***^ |
| ALT (IU/L) |  | 25.39 (15.82) | 25.81 (16.11) | 0.61 |
| LDH (IU/L) |  | 131.49 (25.90) | 136.82 (29.55) | <0.001^***^ |
| GGT (IU/L) |  | 27.05 (29.50) | 34.54 (49.02) | <0.001^***^ |
| AST (IU/L) |  | 25.13 (14.42) | 26.11 (12.97) | 0.17 |
| Albumin (g/dL) |  | 4.22 (0.31) | 4.20 (0.31) | 0.07 |
| Globulin (g/dL) |  | 2.82 (0.42) | 2.89 (0.46) | <0.001^***^ |
| A/G |  | 1.54 (0.28) | 1.49 (0.29) | <0.001^***^ |
| AST/ALT |  | 1.08 (0.31) | 1.10 (0.31) | 0.029^*^ |
| HbA1c (%) |  | 5.54 (0.73) | 6.05 (1.16) | <0.001^***^ |
| Triglycerides (mg/dL) |  | 116.25 (62.51) | 139.39 (74.07) | <0.001^***^ |
| Treat of hypertension | NO | 2616 (77.7%) | 1061 (46.7%) | <0.001^***^ |
|  | YES | 717 (22.3%) | 1337 (53.3%) |  |
| LDL-C (mg/dL) |  | 119.82 (28.34) | 118.06 (32.01) | 0.168 |

Continuous variables are expressed as mean ± standard deviation and compared across groups using the Wald test. Categorical variables are expressed as frequencies (weighted percentages) and compared using the Rao–Scott adjusted chi-square test. DBP, diastolic blood pressure; SBP, systolic blood pressure; HDL-C, high density lipoprotein cholesterol; ALP, alkaline phosphatase; ALT, alanine aminotransferase; LDH, lactate dehydrogenase; GGT, -glutamyl transpeptidase; AST, aspartate transaminase; A/G, albumin / globulin; HbA1c, glycosylated hemoglobin, type A1C; LDL, low density lipoprotein cholesterol. *, P< 0.05; **, P < 0.01; ***, P < 0.001.
